# Predictors of COVID-19 Vaccine Hesitancy in North-Central Nigeria

**DOI:** 10.1101/2023.12.20.23300316

**Authors:** Ayodotun Olutola, Ibrahim Bola Gobir, Deus Bazira, Samson Agboola, Fatimah Ohunene Sanni, Azeez Akanbi Bello, Havilah Onyinyechi Nnadozie, Aisha Adamu, Fatima Bello, Suzzy Angmun Otubo, Mercy Piring’ar Nyang

## Abstract

COVID-19 vaccine hesitancy has emerged as a major challenge to global efforts to control the pandemic, particularly in Nigeria, where hesitancy to other effective vaccines such as polio and measles has been widely reported. Several individual, societal, and structural factors contribute to this behaviour and prevent the effectiveness of COVID-19 prevention efforts.

**Objectives:** This study sought to assess the factors associated with COVID-19 vaccine hesitancy in the six states of north-central Nigeria.

**Methods:** A population-based cross-sectional online survey was conducted among residents using a semi-structured questionnaire adapted from the WHO SAGE vaccine hesitancy scale and distributed via social media networks over 8-weeks.

**Results:** A total of 1,999 responses were received, of which 570 were set aside comprising 512 respondents that resided outside the study area, 12 respondents that reported no knowledge of the COVID-19 vaccine, and 46 entries with missing data. Of 1,429 included in the analysis, 1,008 (70.5%) were willing to be vaccinated and/or already vaccinated and 421 (29.5%) were unwilling to receive the COVID-19 vaccine. Post-secondary education (AOR: 0.51, 0.37-0.69), household income above the minimum wage of 30,000 Naira per month (AOR: 0.68, 0.50-0.94) and people of the Islamic faith (AOR: 0.69, 0.53-0.90) were found to be associated with lower levels of hesitancy. The dominant reasons for hesitancy were concerns about side effects (37.5%), doubt about the existence of COVID-19 (11.0%), and the perception of time required to receive the vaccine (9.6%). Hesitant respondents relied on health workers (33.0%) and social media (23.3%) as their trusted sources of health information, and less than a third (31.4%) followed the advice of their religious and community leaders.

**Conclusion:** All three factors of confidence, complacency and convenience influenced hesitancy in our study. Socioeconomic factors are major drivers of hesitancy. Therefore, hesitancy is as much a social issue as health and requires a multisectoral approach to educating communities and building trust in health and social institutions.

## Introduction

The Coronavirus Disease 2019 (COVID-19) was declared a global pandemic in March 2020. As of August 2023, globally, 769 million cases and 6.9 million deaths were estimated to have occurred. Against higher predictions, Nigeria recorded only 265,000 cases, 266,675and 3,155 deaths from the disease during the same period [1, 2]. The exponential spread of COVID-19 and associated mortality, particularly among the elderly and people with comorbidities called for concerted efforts to develop vaccines. Traditional vaccine development from the preclinical phase to licensing takes on average more than 10 years [3]. However, the 2014 Ebola epidemic spurred the development of novel platforms that shorten the time for sequencing and vaccine trials from years to weeks [4]. Leveraging these platforms and multisectoral collaborations, several vaccines were rapidly produced and approved for distribution. Nearly one year after the first case of COVID-19 was published, Janssen announced the success of a phase 3 trial of a potential COVID-19 vaccine [5]. The bid by developed nations to hasten research and access to vaccines assumed economic and political dimensions and, vaccines were initially more readily available in the developed countries. While nations scrambled for vaccines, a greater challenge was ensuring that eligible populations accepted to be vaccinated.

In March 2021, Nigeria received the first batch of the AstraZeneca vaccine and began vaccinations, prioritizing health workers and individuals most at risk for COVID-19. As of September 21, 2022, 64.36% of the people in North America [6] compared to 15% of the people in Nigeria [7] had been fully vaccinated with COVID-19 vaccine. Several pre-vaccination studies reported varying degrees of reluctance and even refusal to take the vaccine among younger people, people of low socio-economic status, people with a negative attitude towards scientific research and low trust in government and health institutions. Some studies found an association between religious beliefs, history of vaccine hesitancy, history of chronic illnesses, and COVID-19 vaccine hesitancy [8]. Concerns about the side effects of the COVID-19 vaccine were the most frequently expressed reason for the reluctance to COVID-19 vaccines in a study conducted across 5 African countries [9].

The World Health Organization defines vaccine hesitancy as a delay in the acceptance or complete refusal of vaccines despite the availability of vaccine services is regarded as vaccine hesitancy. [10] It has been classified as one of the top ten public health issues. The concept of vaccine hesitancy is complex and context-specific. Factors such as age, culture, socio-economic status, and trust in the healthcare system have been found to influence vaccine hesitancy [11, 12]. The WHO SAGE working group identified three barriers to vaccine uptake known as the 3C’s: Confidence in vaccines and trust in the system that delivers them, complacency, when perceived risks of vaccine-preventable diseases are low or when vaccination is considered secondary to other responsibilities at a given point in time and convenience which denotes the extent to which physical availability, affordability, and access to information and other immunization services exist [13].

Modern anti-vaccine movements are fuelled by claims that vaccines are the root causes of certain diseases. For example, diphtheria, pertussis, and tetanus vaccines were falsely linked to neurological disorders and speculations that measles, rubella, and mumps vaccines were a cause of autism in children [14]. Fears of vaccines containing birth control compounds and a failed meningitis vaccine trial that resulted in catastrophic consequences in Northern Nigeria aggravate tendencies for hesitant behaviour [15, 16]. Other factors such as the unprecedented speed in the development of COVID-19 vaccines, concerns about potential adverse effects, perceived inequities in vaccine distribution, and political inclinations are some contemporary issues fuel COVID-19 vaccine hesitancy. [17]

As vaccines remain the safest strategy for achieving herd immunity, an understanding of the contextual determinants of hesitancy will inform interventions to improve vaccine uptake or alternative interventions to protect hesitant populations.

This study sought to deconstruct the underlying drivers and its correlates including individual, interpersonal, and societal factors influencing COVID-19 vaccine hesitancy among residents in North Central Nigeria.

## Method

### Study design

We obtained data through a population-based cross-sectional online survey conducted among residents in the seven North-central states in Nigeria; Federal Capital Territory, Nasarawa, Niger, Kogi, Benue, Plateau, and Kwara states. The semi-structured survey questionnaire used was adapted from the WHO SAGE vaccine hesitancy scale [13] and refined through a literature review to identify factors critical to vaccine hesitancy. The survey was developed using REDCap and was distributed via social media networks (WhatsApp, Telegram, LinkedIn, Instagram, and Facebook) over 8 weeks from the 7^th^ of March 2022 to the 30^th^ of April 2022.

### Data Collection

The convenience sampling method was used to recruit respondents for the study. The entry page of the survey contained survey information and study objectives. Respondents’ consent was documented by clicking on the agreement button after reading the survey information. The respondents completed a questionnaire in four sections that captured socio-demographic characteristics, information about acceptance of COVID-19 vaccine and COVID-19-related perceptions. Approval was received from the Nigerian National Health Research Ethics Committee and Georgetown University Institutional Review Board.

### Inclusion and Exclusion Criteria

1,487 persons who met the inclusion criteria to participate in the study were recruited through distribution on social networks such as private messaging, electronic mails, social media platforms such as WhatsApp, LinkedIn, Instagram, and Facebook.

Exclusion criteria include age below 18 years or residence outside of the study area.

All participants provided written and informed permission.

### Sample Size Determination

The sample size calculation is based on the adult population in Nigeria, which is 115,897,765 (Nigeria Bureau of Statistics). with a 95% confidence interval and a margin of error of 2.5% with an estimated retention rate of 80% using StatCalc. The calculated minimum sample size was 1,771 for persons older than 18 years living in North central Nigeria. A total of 1,999 responses were received from the distributed questionnaires through distribution on social media platforms such as WhatsApp, LinkedIn, Instagram and Facebook.

### Statistical Analysis

The summary statistics were obtained for all variables using descriptive statistics including frequencies, percentages, means, and standard deviations. A chi-square test was used to examine the association of sociodemographic characteristics with attitudes toward COVID-19 Vaccination. The analysis to examine the factors associated with vaccine hesitancy was carried out using logistic regression. All analysis was performed at a 5% significance level and carried out using IBM Statistical Product and Service Solutions (SPSS) software, version 25.

### ETHICAL CONSIDERATIONS

#### Informed consent

The survey’s home page contained information about the study’s objectives, eligibility criteria, data protection, and researcher disclaimers. Participants provided written informed consent. It was deemed sufficient to provide informed consent if a participant checked the “I agree” box on the survey. Participants’ entries that did not match the inclusion criteria were not processed for data analysis.

#### Confidentiality

Each submission was made anonymously. Participants’ personal information was not gathered. The subjects’ identities were kept hidden.

#### Risks and benefits

There were no negative consequences for the subjects’ rights or well-being. Participating in the study provided no immediate benefits.

#### Ethical clearance

The study protocol was submitted for assessment and approval to the Nigerian Health Research Ethics Committee (NHREC).

## Results

A total of 1,999 survey entries were recorded and 570 were excluded comprising 512 respondents that resided outside the study area, 12 respondents that reported no knowledge of the COVID-19 vaccine, and 46 entries with missing data. Among the 1,429 respondents in the study, 1,008 (70.5%) were willing to accept or had received at least one dose of the COVID-19 vaccine while 421 (29.5%) were unvaccinated and hesitant.

**Fig 1.**
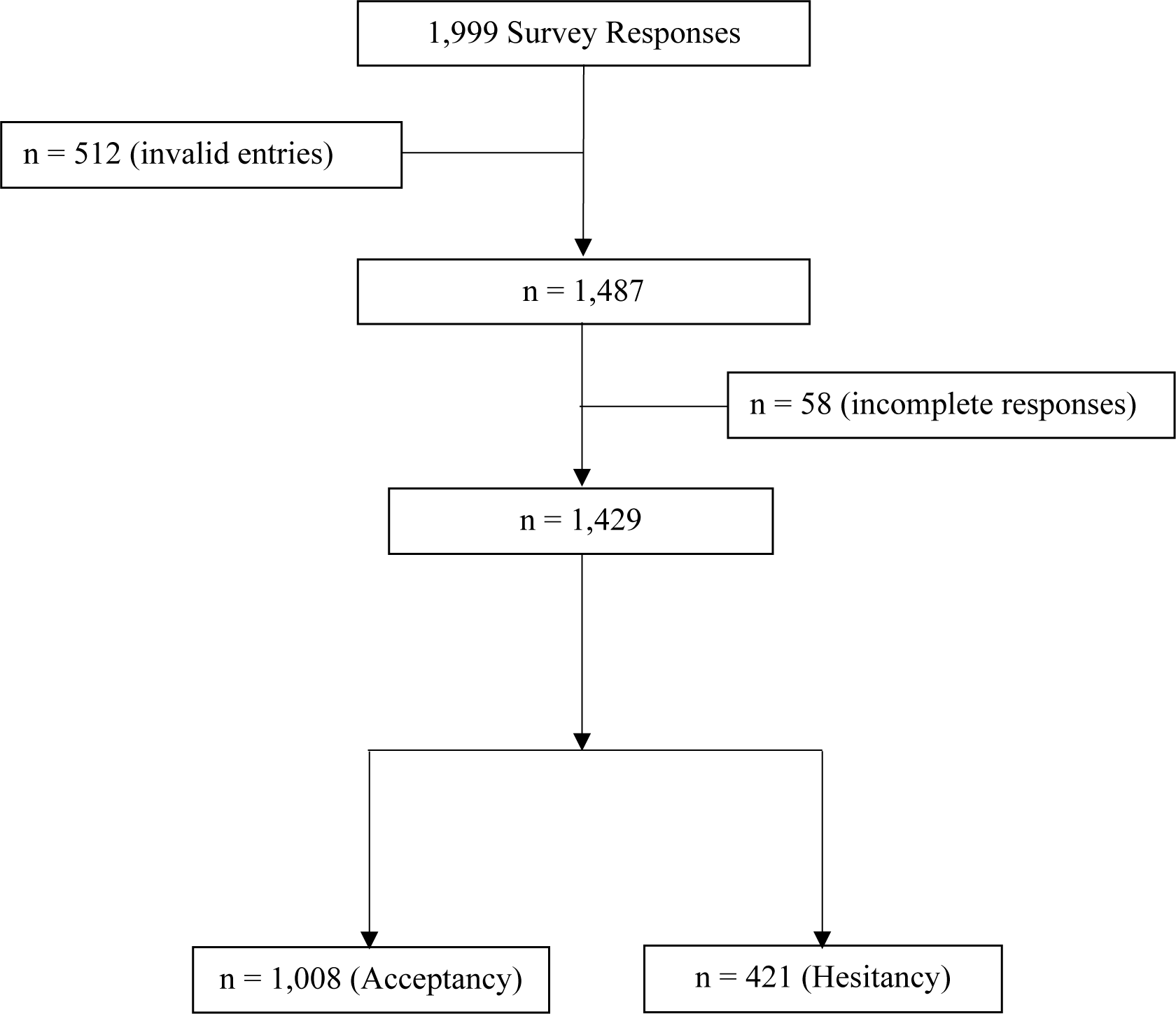
Flow diagram of the study sample size.

### Demographic characteristics of the respondents

More than third (36.5%) of the respondents were residents of the FCT while the rest resided in Benue (5.9%), Kogi (10.4%), Kwara (3.3%), Nasarawa (15.4%), Niger (16.6%), and Plateau States (11.8%). The majority of respondents were male (60.3%), 18 -25yrs old (43.9%), single (64.6%), Christian faith (57.4%), post-secondary-education (80.3%) and employed (32.4%). Close to half of the respondents (45.7%) had a monthly income between N30,000 and N150,000. About two-thirds of the respondents (68.2%) reported losing their income during the COVID-19 pandemic.

**Table 1.**
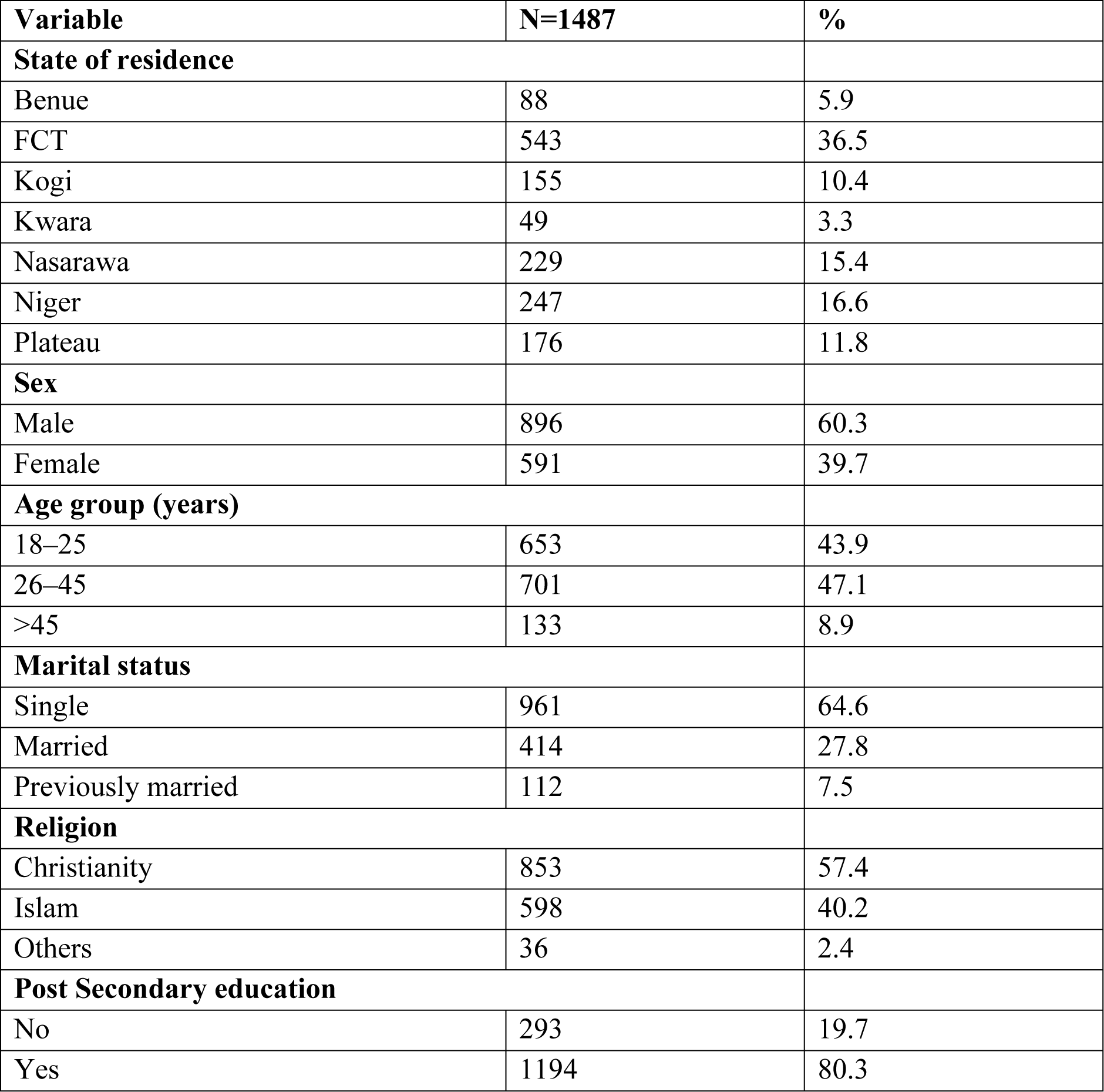
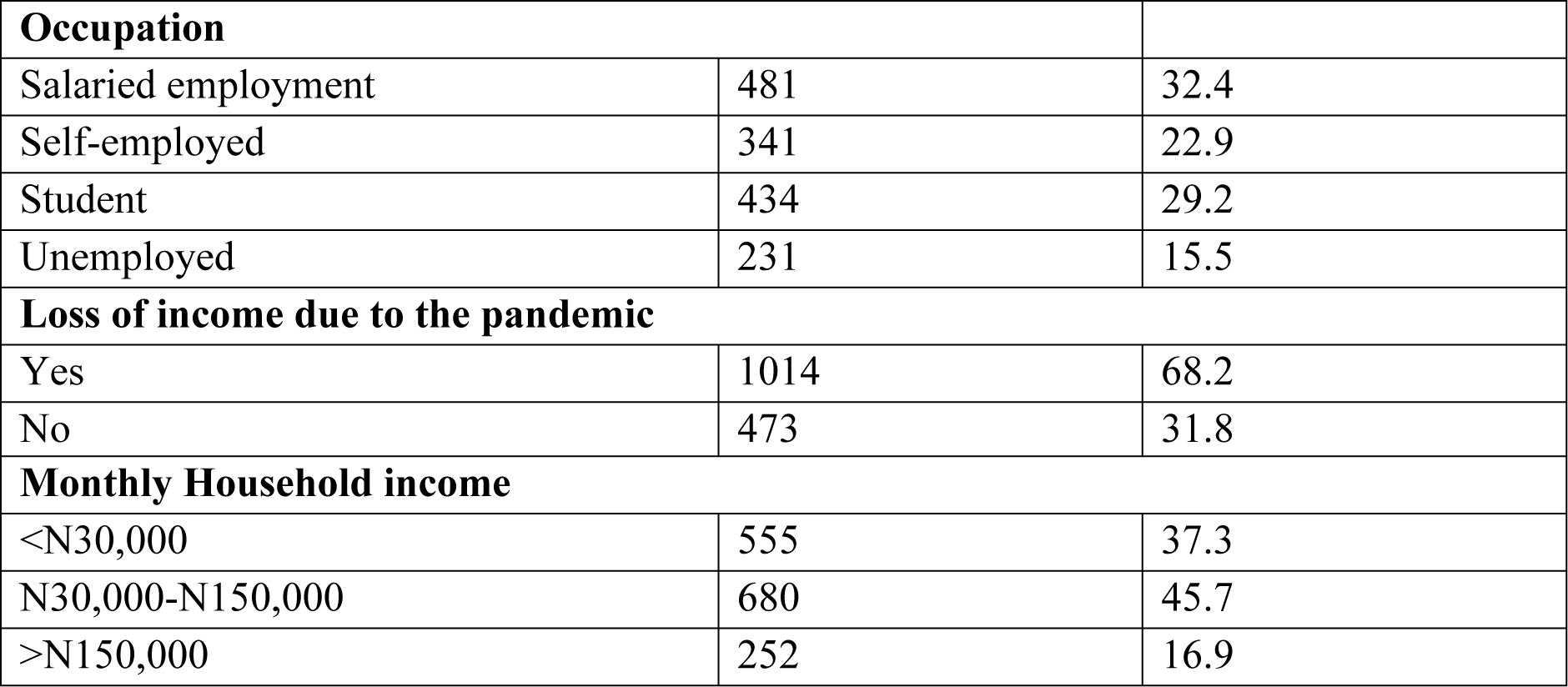
Demographic characteristics of all respondents, N=1,487.

### Sociodemographic predictors of COVID-19 Vaccine hesitancy

Persons with post-secondary education were 50% less likely to be hesitant than those without post-secondary education while persons of the Muslim faith were 30% less likely to be hesitant compared to the Christian faith. Odds for hesitancy were increasingly reduced for income bands above 30,000 Naira.

**Table 2.** Sociodemographic predictors of COVID-19 vaccine hesitancy among respondents.

| Variable | Total N | Hesitant n (%) | P Value | Crude OR (95% CI) | Adjusted OR (95% CI)* | P value** |
| --- | --- | --- | --- | --- | --- | --- |
| **Total** | 1,429 | 421 (29.5) |  |  |  |  |
| **State** |  |  | 0.12 |  |  |  |
| FCT | 524 | 149 (28.4) |  | Ref | Ref | 0.28 |
| Benue | 79 | 28 (35.4) |  | 1.38 (0.84-2.27) | 1.17 (0.69-1.97) |  |
| Kogi | 147 | 53 (36.1) |  | 1.42 (0.96-2.09) | 1.20 (0.80-1.80) |  |
| Kwara | 46 | 11 (23.9) |  | 0.79 (0.39-1.60) | 0.80 (0.39-1.64) |  |
| Nasarawa | 223 | 52 (23.3) |  | 0.77 (0.53-1.10) | 0.70 (0.48-1.03) |  |
| Niger | 238 | 75 (31.5) |  | 1.16 (0.83-1.62) | 1.12 (0.78-1.61) |  |
| Plateau | 172 | 53 (30.8) |  | 1.12 (0.77-1.63) | 1.03 (0.70-1.53) |  |
| **Sex** |  |  | 0.52 |  |  |  |
| Female | 562 | 171 (30.4) |  | Ref | Ref | 0.96 |
| Male | 867 | 250 (28.8) |  | 0.93 (0.73-1.17) | 1.01 (0.79-1.29) |  |
| **Age** |  |  | 0.31 |  |  |  |
| 18-25 | 618 | 195 (31.6) |  | Ref | Ref | 0.53 |
| 26-45 | 678 | 190 (28.0) |  | 0.84 (0.67-1.05) | 0.85 (0.63-1.15) |  |
| >45 | 133 | 36 (27.1) |  | 0.81(0.53-1.22) | 0.78 (0.44-1.37) |  |
| **Marital status** |  |  | 0.25 |  |  |  |
| Single | 916 | 274 (29.9) |  | Ref | Ref | 0.43 |
| Married | 404 | 109 (27.0) |  | 0.87 (0.67-1.12) | 0.94 (0.68-1.30) |  |
| Previously married | 109 | 38 (34.9) |  | 1.25 (0.83-1.91) | 1.31 (0.77-2.33) |  |
| **Religion** |  |  | 0.005 |  |  |  |
| Christianity | 817 | 258 (31.6) |  | Ref | Ref | 0.005 |
| Islam | 577 | 147 (25.5) |  | 0.74 (0.58-0.94) | 0.69 (0.53-0.90) |  |
| Others | 35 | 16 (0.83-1.9) (45.7) |  | 1.82 (0.92-3.61) | 1.62 (0.80-3.30) | <0.001 |
| **Post Secondary education** |  |  | <0.001 |  |  |  |
| No | 285 | 124 (44.0) |  | Ref | Ref | <0.001 |
| Yes | 1,144 | 297 (26.0) |  | 0.46 (0.35-0.60) | 0.51 (0.37-0.69) |  |
| **Occupation** |  |  | 0.002 |  |  |  |
| Self-employed | 330 | 119 (36.1) |  | Ref | Ref |  |
| Salaried worker | 471 | 115 (24.4) |  | 0.57 (0.42-0.78) | 0.77 (0.55-1.08) | <0.001 |
| Student | 404 | 112 (27.7) |  | 0.68 (0.50-0.93) | 0.60 (0.41-0.85) |  |
| Unemployed | 224 | 75 (33.5) |  | 0.89 (0.62-1.28) | 0.67 (0.45-1.02) |  |
| **Monthly Household income** |  |  | 0.001 |  |  |  |
| <N30,000 | 523 | 185 (35.4) |  | Ref | Ref | 0.03 |
| N30,000-N150,000 | 660 | 177 (26.8) |  | 0.67 (0.52-0.86) | 0.68 (0.50-0.94) |  |
| >N150,000 | 246 | 59 (24.0) |  | 0.58 (0.41-0.81) | 0.58 (0.37-0.92) |  |

### Other factors influencing Vaccine Hesitancy

Concerns about side effects were the most common reasons for vaccine hesitancy (37.5%). Others expressed reasons such as doubt about the existence of COVID-19 (11.0%), the perception of time required to receive the vaccine (9.6%), dislike or fear of needles (7.2%) and possible complications caused by underlying medical conditions (5.5%). A few believed that the vaccines were not effective (6.5%) (Fig. 2).

**Fig 2.**
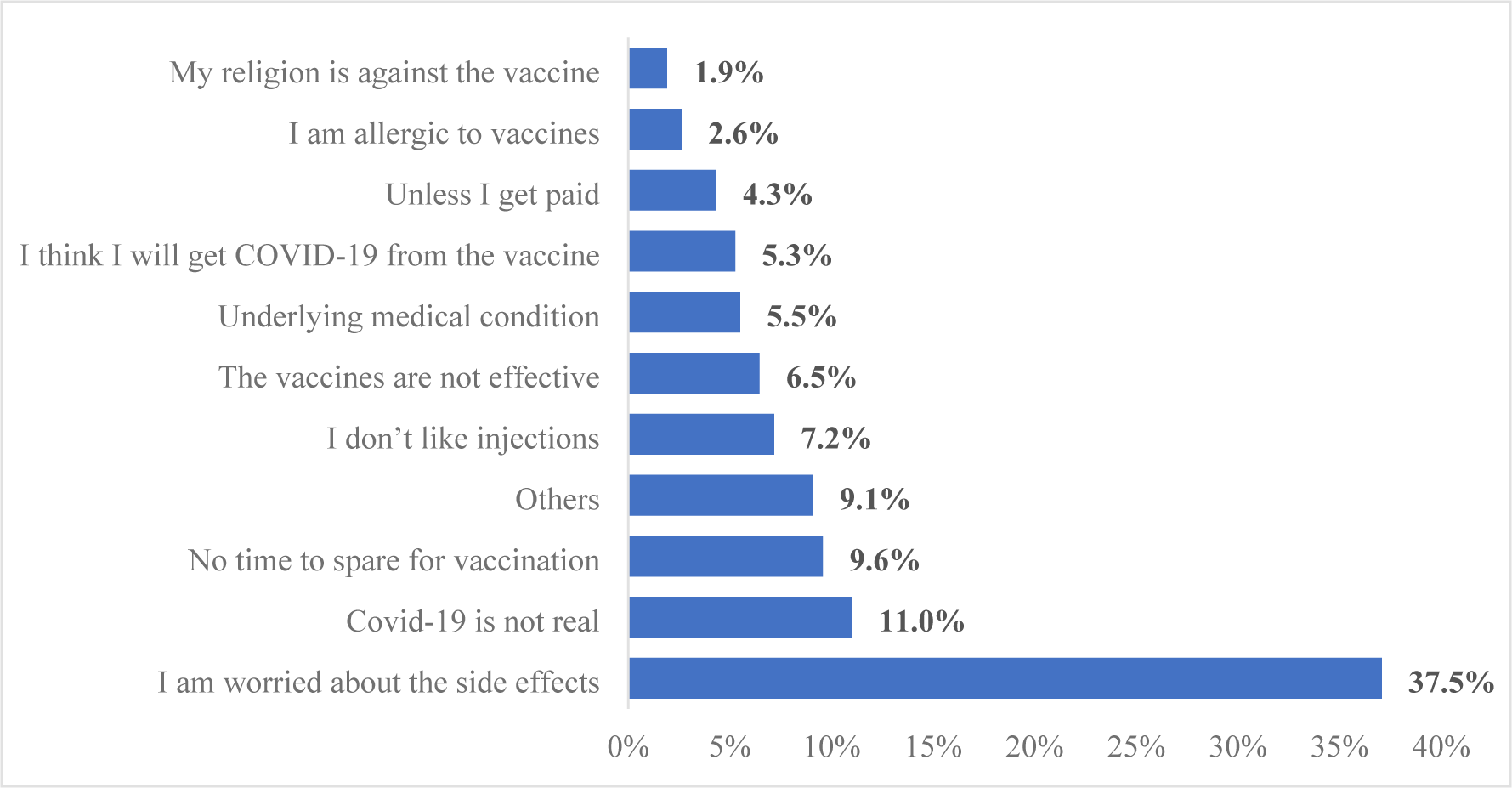
Other factors influencing Vaccine Hesitancy.

More than 70% of respondents expressed lack of trust in government to purchase the highest quality of vaccine or in the pharmaceuticals to produce safe vaccines. Over 60% thought the vaccine to be unsafe, non-essential and the vaccination process to be inconvenient. The majority held the opinion that non-pharmaceutical interventions are sufficient protection against and can eradicate COVID-19 The majority of community leaders including religious leaders (85.6%) support vaccination, however 68.8% hesitant expressed unwillingness to follow the advice of these leaders. (Table 3).

**Table 3.** COVID-19 vaccine-related perceived beliefs, concerns, risk, trust, and location perceptions.

| Factors | Yes |  | No |  |
| --- | --- | --- | --- | --- |
| Leaders, gatekeepers, and pro-vaccination |  |  |  |  |
| Do religious leaders in your community support taking the COVID-19 vaccine? | 360 | 85.5% | 61 | 14.5% |
| Do politicians, teachers, and health workers in your community support vaccination? | 392 | 93.1% | 29 | 6.9% |
| Do you follow the advice of your religious/cultural leaders about the COVID-19 vaccination? | 132 | 31.4% | 289 | 68.6% |
| Does your religion/culture recommend against the COVID-19 vaccination? | 66 | 15.7% | 355 | 84.3% |
| **Political influences** |  |  |  |  |
| Do you trust that your government is deciding in your best interest concerning the COVID-19 vaccine | 156 | 37.1% | 265 | 62.9% |
| Are you convinced that your government purchases the highest quality of the COVID-19 vaccine | 120 | 28.5% | 301 | 71.5% |
| Do you trust that the government is making the best efforts to store the vaccines in the right conditions? | 133 | 31.6% | 288 | 68.4% |
| **Geographical barriers** |  |  |  |  |
| Does distance limit you from getting the vaccine? | 157 | 37.3% | 264 | 62.7% |
| Does clinic time or waiting at the clinic prevent you from getting the COVID-19 vaccine? | 182 | 43.2% | 239 | 56.8% |
| **Pharmaceutical industry** |  |  |  |  |
| Do you think governments are pushed by the pharmaceutical industries to recommend vaccines? | 315 | 74.8% | 106 | 25.2% |
| Do you trust the pharmaceutical companies that produce the COVID-19 vaccine? | 118 | 28.0% | 303 | 72.0% |
| **The Vaccines/Vaccination issues** |  |  |  |  |
| Do you think the vaccine is safe | 142 | 33.7% | 279 | 66.3% |
| The COVID-19 vaccine is essential for us | 160 | 38.0% | 261 | 62.0% |
| Do you trust the healthcare workers to safely administer the vaccine to you? | 200 | 47.5% | 221 | 52.5% |
| Is the vaccination process convenient, i.e., is it easier for you to get vaccinated | 159 | 37.8% | 262 | 62.2% |
| Do you think that if everyone in society maintains preventive measures (face masks, social distancing, etc.), COVID-19 can be eradicated without vaccination? | 285 | 67.7% | 136 | 32.3% |

The majority (33.0%) of the respondents indicated that health workers were their most trusted source of information. Social media was trusted most by 24.0% of the respondents and 23.3% trusted family, friends, and community while 14.7% indicated mass media as the most trusted source of information (Fig 3).

**Fig. 3.**
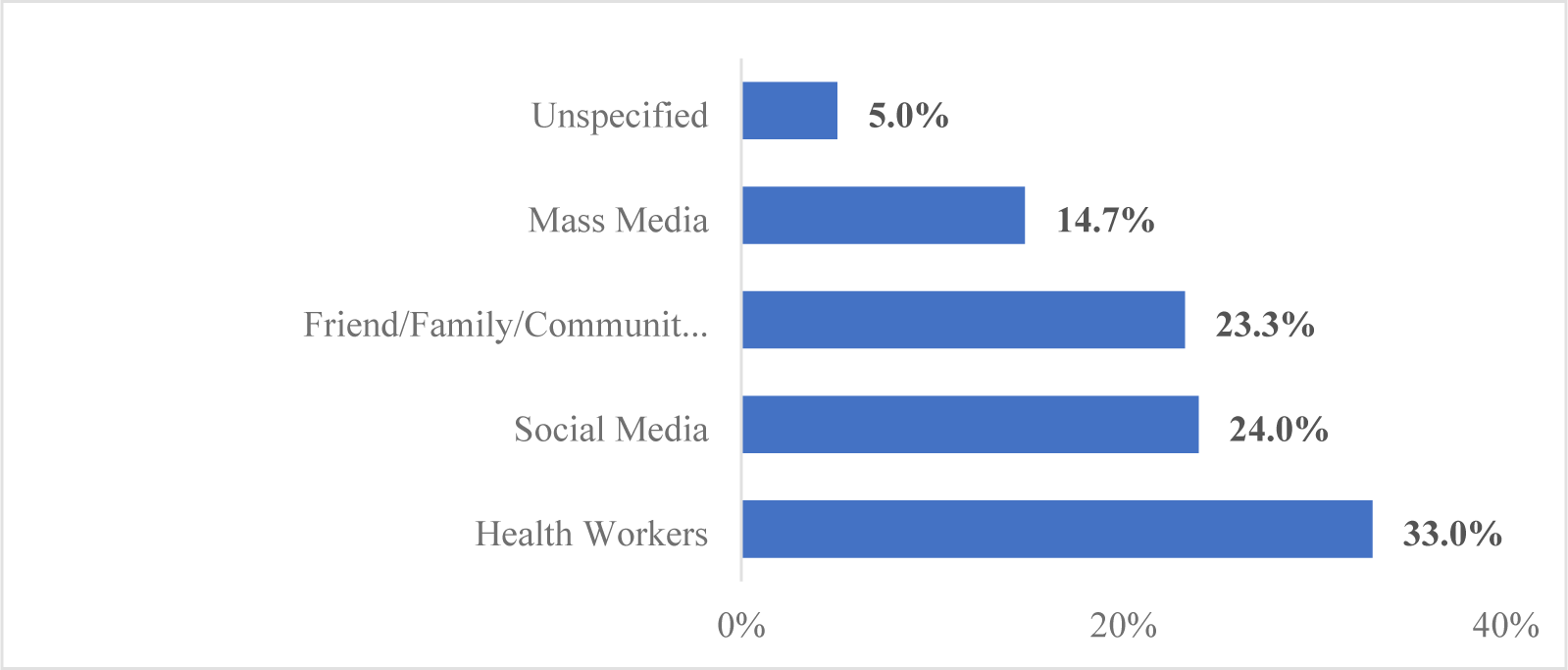
The most trusted source of information about the vaccine.

## Discussion

In this study, 29.3% of the respondents were hesitant to receive the COVID-19 vaccine. A systematic review of COVID-19 vaccine hesitancy rates in Africa reported that vaccine hesitancy rates varied between 2.1% to 93.1% based on studies from 17 African countries [18]. Hesitancy rates above 50% have also been reported in China, Malaysia, the U.S.A., Australia, Pakistan and Italy [11, 19, 20, 21]

Some sociodemographic characteristics such as religious status, education and employment status were found to be significantly associated with vaccine hesitancy in the study. Post-secondary education was associated with a 50% risk reduction in the rate of hesitancy. This is consistent with most other studies with similar findings reinforcing the value of education in influencing health-seeking behaviours. [22]. Access to correct information about the vaccine and peer pressure in schools and workplaces contribute to knowledge and attitudes towards health interventions. [23] Students had a lower risk of hesitancy compared to self-employed persons. Increasing household income above the national minimum wage of 30,000 was significantly associated with a lower risk of hesitancy, with respondents earning over 150,000 monthly demonstrating approximately a 50% reduction in risk. Muslim faith was found to be associated with a 50% lower risk of hesitancy compared to the Christians. This may not be un-associated with the anti-vaccine messaging that emanated from some prominent Christian leaders at the beginning of the pandemic. [24] Hesitancy rates were comparable among different gender and age categories.

In this study, vaccine hesitancy was influenced by a combination of confidence, convenience, and complacency factors.

Lack of confidence was demonstrated in the belief about the safety of the vaccine, and the risk of side effects. This ranked as the highest concern among others, as nearly half of the vaccine-hesitant respondents cited concerns about the safety and side effects of the vaccine, and this aligns with findings from other studies [25, 26, 27]. They also expressed a lack of trust in both the government of Nigeria and pharmaceutical companies to deliver the vaccine with the sole purpose of improving quality of life. Similar attitudes to vaccines have been demonstrated in other settings such as polio vaccination drive where citizens in Northern Nigeria perceived the vaccines to contain compounds with sterilization properties. [28]. The unprecedented speed of production of these vaccines and the global issues that limited access to specific types of vaccines were diversely interpreted by local opinion leaders and the citizenry. The Astra Zeneca vaccine was quickly tagged as inferior to the preferred options in the USA such as Moderna and Pfizer vaccines. These findings are consistent with the results of other COVID-19 acceptance studies as well as the broader vaccine hesitancy literature. According to Larson et al in 2018 [24], a lack of trust in the broader society, including government, economic and health systems is directly or indirectly related to vaccination and can have a profound effect on the trust in vaccines.

Complacency factors were demonstrated by those who expressed that the COVID infection was not real, the vaccine was not required and those who felt the use of universal precautions and personal protective equipment were adequate to address the epidemic. Similar views have been documented elsewhere, with the belief that COVID-19 was a politically or business-motivated construct for selfish reasons. Among others, some rumoured that it was engineered to facilitate the inoculation of tracking chips into humans or a ploy by pharmaceutical companies to sell their products. Some others voiced COVID-19 as a punishment from God, or an effect of 5G technology deployment, while others believed it was just the regular flu [29]. A low level of education and exposure to health information may have facilitated the spread of misinformation and reduced trust in the vaccines and their efficacy. Vaccine-hesitant respondents indicated that health workers and the social media were their most trusted source of health information. they believed the media reports claiming the vaccine was not safe as other research around vaccine hesitancy has reported similar findings. [22, 26] Public health messaging has traditionally relied on publication in academic journals, media campaigns on television and radio, or visually appealing infographics. Research has shown that healthcare providers and pro-vaccine groups are not as active and connected as antivaccine movements on social media. Concern for damage to professional image, data security issues and the risk of perpetuating personal biases have been identified as reasons for the limited use of social media by healthcare providers [30]. This is because many conspiracy theories and propaganda about the COVID-19 vaccine originate and spread on social media platforms.

Convenience factors were dominated by factors that affect access to the vaccine such as the distance from homes to vaccination centres and the perceived waiting time required to complete the vaccination. All these factors relate to both financial and non-financial costs of receiving the vaccines. Factors such as the low level of income of most respondents could contribute to the unaffordable cost and convenience issues.

We acknowledge some limitations of the study which may have affected the generalizability of the study. Completion of the survey was based on self-selection and may have introduced some bias in the study population. The online nature of the survey may have led to the underrepresentation of people with limited access to mobile devices and may have influenced the demographic and socioeconomic characteristics of the respondents with the recruitment of more of the younger population below 35 years and more educated populations. In addition, the inferences from this study relate to the COVID-19 vaccine and may not apply to vaccines for other diseases.

## Conclusion

The study demonstrates the impact of social determinants of health on the acceptance and uptake of COVID vaccine. Socioeconomic factors such as educational attainment, income and faith are major drivers of hesitancy. These and other findings influenced varied degrees of confidence, complacency and convenience factors that were associated with hesitancy in the study. Hesitancy is as much a social issue as health and requires a multisectoral approach in educating communities and building trust in health and social institutions. Approaches to health education must be adaptive to build societal trust in medical products and service providers using conventional platforms such as educational institutions, congregational settings, media, and the social media to disseminate health messages and scientific breakthroughs. Such interventions will be more effective when routinized prior to major events like the COVID-19 pandemic.

## Data Availability

Data set supporting the findings of this study are available within the paper and its supporting files (Supplementary data)

## Acknowledgement

We would like to express gratitude to Center for Global Health Practice and Impact (CGHPI), Center for Clinical Care and Clinical Research (CCCRN), Georgetown Global Health Nigeria (GGHN), Savannah Health System Innovation Ltd (SHSIL), and everyone who supported and contributed to protocol development, data collection and development of the manuscript.

## Funding

The study was funded by Georgetown University Medical Center, Dean of Research. However, the funder did not play any role in the research.

## Notes

### Competing Interest Statement

The authors have declared no competing interest.

### Funding Statement

Yes

### Author Declarations

Approval was received from the Nigerian National Health Research Ethics Committee and Georgetown University Institutional Review Board.

